# Assessing the Impact of the Inflation Reduction Act on Medicare Prescription Drug Coverage

**DOI:** 10.1101/2025.07.24.25332155

**Authors:** Yi Wolf Zhang, Michael G. Blyumin, Cori Qu, Xu Guo, Minlu Zhang

**Author notes:** These authors contributed equally to this work (Yi Wolf Zhang, Michael G. Blyumin).

## Abstract

**Importance:** The Inflation Reduction Act (IRA) introduced significant reforms to Medicare Part D in 2025, aiming to reduce medication costs for beneficiaries by eliminating the coverage gap (“donut hole”) with a $2,000 annual out-of-pocket cap. However, insurers’ responses to these policy reforms—and their potential implications for medication affordability and access—have not been systematically evaluated.

**Objective:** To assess how Medicare Part D plans adjusted benefit design and formulary structures in response to the IRA and determine the impact of these adjustments on beneficiaries’ financial responsibilities.

**Design, Setting, and Participants:** Retrospective comparative analysis of Medicare Part D plans (Standalone Prescription Drug Plans [PDPs] and Medicare Advantage Prescription Drug [MAPD] plans), comparing plan structure, formulary coverage, utilization management, and cost-sharing details from January 2024 to January 2025. Analyses were conducted using publicly available Centers for Medicare & Medicaid Services (CMS) datasets.

**Main Outcome(s) and Measure(s):** Primary outcomes included changes in annual deductible amounts, cost-sharing structures (co-payment vs co-insurance), formulary coverage and utilization management practices, and patient out-of-pocket costs for GLP-1 medications, including first-fill cost burden.

**Results:** Across PDPs, mean annual deductibles significantly increased from $384.7 (95% CI, 369.2–400.1) to $454.0 (95% CI, 432.9–475.1); MAPD plans showed a greater increase from $98.7 (95% CI, 93.6–103.8) to $249.0 (95% CI, 241.4–256.6). In MAPD plans, co-insurance style coverage utilization drastically increased for tier 3 medications (2024: 6.3%, 2025: 38.1%). Specifically, for GLP-1s, while overall coverage decreased, preferred drugs like Ozempic and Mounjaro experienced expanded coverage. However, first-fill out-of-pocket expenses increased substantially due to higher deductibles and increased costs associated with co-insurance style coverage. First-fill costs exceeding $600 rose from 40%-45% (2024) to over 80% (2025) in PDPs and from less than 1% to approximately 13% in MAPD plans.

**Conclusions and Relevance:** Medicare Part D plans in 2025 were strategically designed to increase beneficiaries’ financial responsibilities via higher deductibles, increased co-insurance cost-sharing, and restricted formulary coverage. While overall annual patient medication cost burdens will decrease, the cost of filling each medication will increase, which may negatively impact medication access and adherence.

**Key Points:** *Question:* How did 2025 Medicare Part D plans respond and adapt to new IRA policies, and how is this affecting medication affordability and access?

*Findings:* Part D plans have adopted higher deductibles, shifted from co-payment to co-insurance, and limited formulary coverage, which has increased beneficiaries’ financial responsibility and negatively impacted medication access.

*Meaning:* Although IRA provisions may reduce patients’ annual medication out-of-pocket costs, Part D plans are adopting new strategies that increase patients’ financial challenges.

## Introduction

Medicare provides health insurance for individuals aged 65 and older and younger individuals with disabilities. As of 2025, 68.5 million individuals are enrolled in Medicare, and 81.3% have Medicare Part D coverage.^1^ Medicare beneficiaries obtain prescription coverage either through Standalone Prescription Drug Plans (PDPs), supplementing traditional Medicare (Part A and B), or via Medicare Advantage Prescription Drug (MAPD) plans, which combine medical and prescription benefits.^2^ Despite existing coverage, many Medicare beneficiaries face significant challenges due to rising prescription drug costs, particularly for chronic conditions requiring long-term brand-name medications.^3–7^

To reduce prescription drug costs for Medicare beneficiaries, beginning in 2025, the Inflation Reduction Act (IRA) introduced several provisions to Part D: eliminating the coverage gap (donut hole), establishing the Drug Manufacturers Discount Program, implementing a new $2,000 annual out-of-pocket threshold, and introducing the Medicare Prescription Payment Plan (MPPP).^8^ These changes were intended to reduce out-of-pocket costs and enhance medication access and adherence.^9–11^

Part D plans are designed and administered by third-party insurers within the Centers for Medicare & Medicaid Services (CMS) guidelines.^12^ These insurers have considerable flexibility in structuring their plans by adjusting premiums, deductibles, cost-sharing approaches (co-payment or co-insurance), drug-tier classifications, and utilization management practices such as Prior Authorization (PA).^13,14^

Under the IRA, Medicare prescription coverage has three phases: deductible, initial coverage, and catastrophic coverage. In 2025, the standard maximum deductible is $590. The deductible must be fully paid before medications are covered under the initial coverage phase. Initial coverage is provided via co-insurance or co-payment style coverage. When beneficiaries incur $2,000 in True Out-of-Pocket (TrOOP) medication costs, they transition into catastrophic coverage, where they are no longer required to share costs for covered medications (eFigure 1).

**Figure 1.**
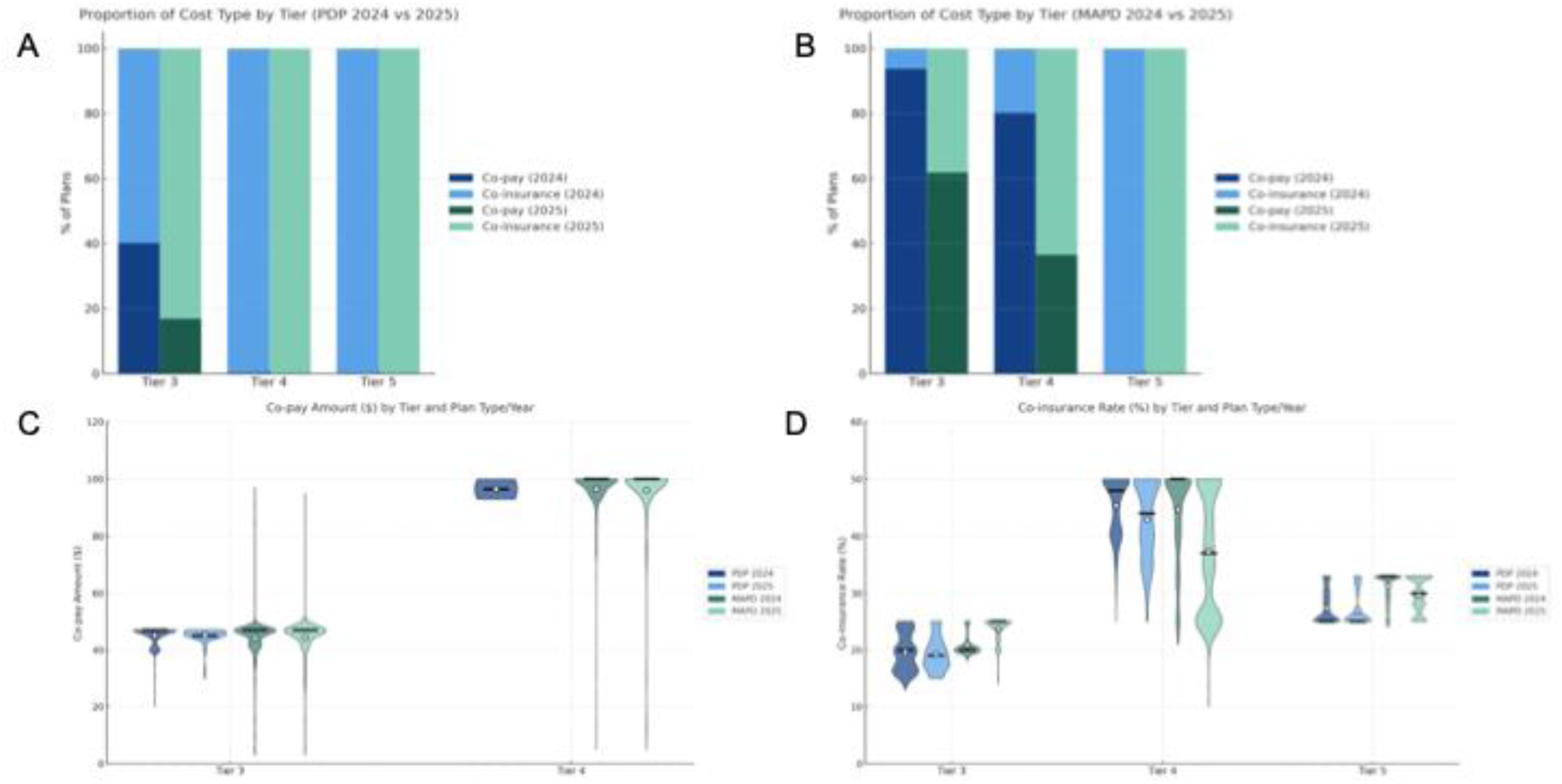
Changes in Medicare Part D Cost-Sharing Structures, 2024 vs 2025. **(A)** Distribution of cost-sharing types among PDPs for high-cost tiers (Tiers 3–5) in 2024 and 2025. **(B)** Distribution of cost-sharing types among MAPD plans for high-cost tiers (Tiers 3–5) in 2024 and 2025. **(C)** Co-payment amount distributions by tier for PDP and MAPD plans in 2024 and 2025. **(D)** Co-insurance rate distributions by tier for PDP and MAPD plans in 2024 and 2025. Black bars represent the median and white dots represent the mean.

Co-insurance and co-payment are cost-sharing methods for beneficiaries’ out-of-pocket (OOP) expenses.^8^ Co-payments are defined as a fixed cost for all medications within a specific tier, while co-insurance is a fixed percentage of the retail price for all medications in the same tier. For example, a tier 3 co-payment is commonly $47 for a 30-day supply, while co-insurance ranges from 15% to 25%. Assuming a brand-name medication list price of $500 per 30 days, 25% co-insurance results in a $125 OOP cost, whereas the co-payment model would keep the $47 cost, regardless of the retail price. Thus, co-insurance generally imposes a greater financial burden on beneficiaries.^15^ This cost discrepancy between co-payment and co-insurance becomes more apparent as the retail cost increases.^16^

How insurance companies respond to these significant IRA-driven policy changes has not been comprehensively evaluated. Thus, the subsequent implications on medication affordability and access are not well understood.^9,17^ Here, we systematically compare Medicare prescription coverage in 2025 versus 2024 (pre-IRA policies) to characterize insurers’ responses and their potential impact on beneficiaries’ ability to afford critical prescription medications.

## Methods

Data from the Centers for Medicare & Medicaid Services (CMS) Public Use Files was utilized to analyze plan information, benefit designs, and formulary coverage (January 2024 and January 2025). The analysis included PDPs and MAPD plans, while Special Needs Plans (SNPs) and Medicaid-Medicare plans were excluded. Enrollment data from the CMS Monthly Report files for March 2024 and March 2025 were used to examine specific enrollment numbers. The average retail price of GLP-1 medications was calculated based on the average list price across all plans. All analyses were performed using Python version 3.9.13. This study is exempt from institutional review board (IRB) review since only publicly available data was used. EQUATOR guidelines were followed for all reporting.

## Results

### Changes in Part D Deductibles and Premiums

Following changes to Medicare Part D policy, annual deductibles for both PDPs and MAPD plans increased significantly from 2024 to 2025 (Table 1). Among PDPs, the mean annual deductible increased from $384.7 (95% CI, $369.2 – $400.1) in 2024 to $454.0 (95% CI, $432.9 – $475.1; p <0.0001) in 2025. Additionally, the proportion of PDPs with a full deductible increased from 55.5% to 66.9%, reflecting a strategic shift toward maximizing patient OOP costs before plan coverage begins.

**Table 1.**
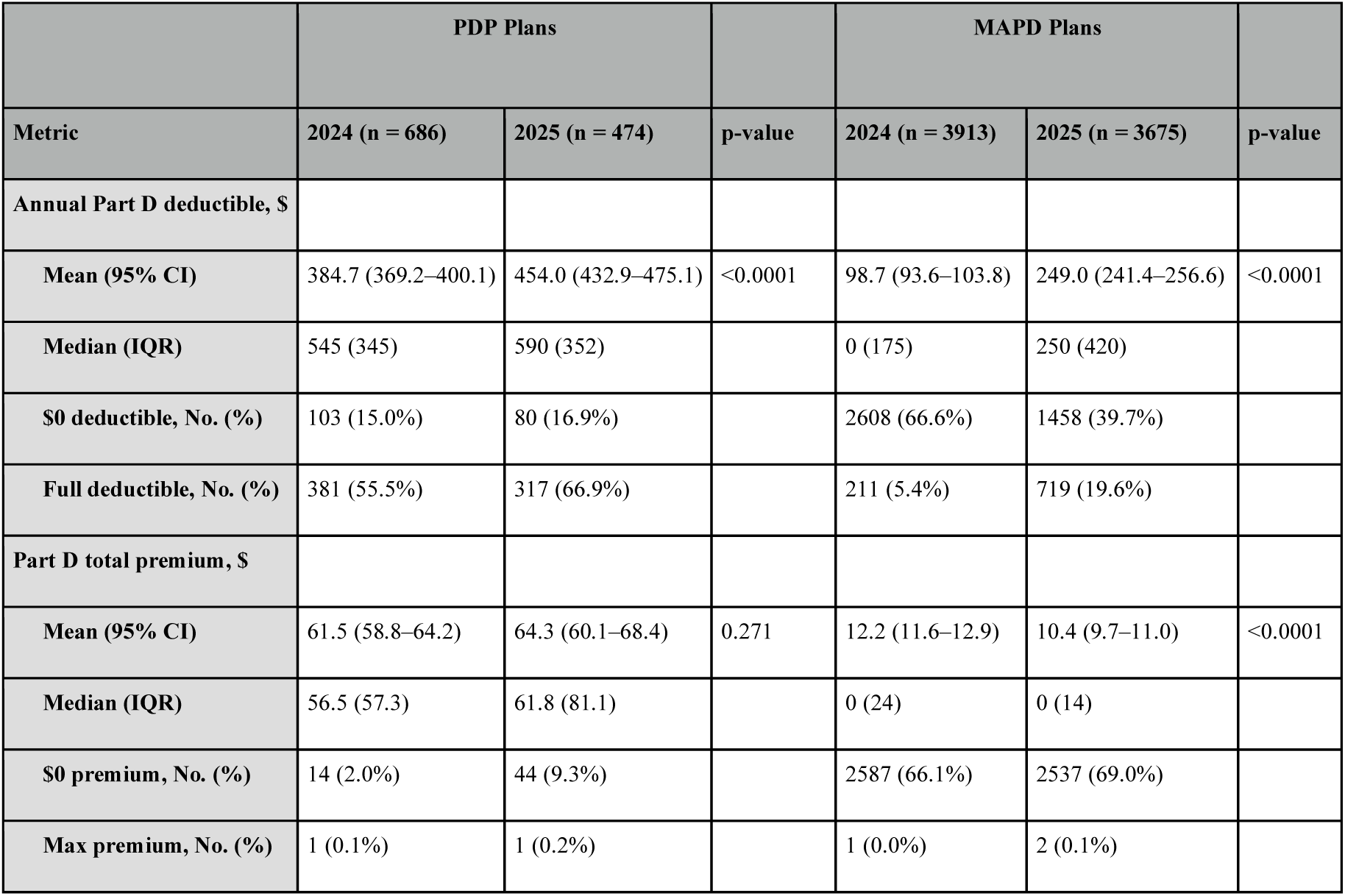
Changes in Medicare Part D Deductibles and Premiums, 2024 vs 2025.

MAPD plans experienced an even greater relative increase, with mean deductibles climbing approximately 2.5-fold, from $98.7 (95% CI, $93.6–$103.8) in 2024 to $249.0 (95% CI, $241.4–$256.6; P < .0001) in 2025. The median deductible increased substantially from $0 (IQR: $175) in 2024 to $250 (IQR: $420) in 2025. The proportion of MAPD plans with $0 deductible dropped from 66.6% in 2024 to 39.7% in 2025, while those with full deductible rose from 5.4% to 19.6%. These trends indicate that MAPD plans have shifted more upfront prescription costs directly to beneficiaries.

Despite substantial changes in deductibles, there was no significant difference in premiums for PDPs between 2024 and 2025, while MAPD plan premiums modestly decreased from an average of $12.2 in 2024 to $10.4 in 2025, further emphasizing insurers’ focus on deductible adjustments as the primary mechanism for cost management.

### Changes in Benefit Design (Co-payment vs Co-insurance)

To understand how Part D plans restructured beneficiaries’ cost-sharing, we analyzed shifts between fixed-dollar co-payments and percentage-based co-insurance across Tier 3 (preferred brand-name medications) and Tier 4-5 medications (non-preferred/specialty medications; Figure 1, eTable 1).

The majority of PDPs consistently use co-insurance coverage for Tier 4 and 5 medications. However, for Tier 3 medications, we observed a significant decrease in co-payment coverage from 40.2% to 16.9% from 2024 to 2025. As a result, 83.1% of 2025 PDP plans employed co-insurance coverage, indicating an increase in beneficiaries’ medication fill costs compared to 2024.

More interestingly, 93.6% of MAPD plans used co-payment for initial coverage of Tier 3 medications in 2024, which dropped drastically to 61.7% in 2025. Similarly, Tier 4 co-payment dropped sharply from 80.1% in 2024 to 36.5% in 2025. This indicates a marked shift toward the co-insurance coverage strategy to allocate more financial responsibility to patients (within the $2000 TrOOP cap). Many MAPD plan beneficiaries who previously benefited from predictable co-payments are now exposed to greater financial uncertainty and hardship as they face co-insurance coverage for the first time.

No significant difference was found for the exact co-payment amount for initial coverage of Tier 3 and 4 medications between 2024 and 2025 (Figure 1C). However, for Tier 3 medications under MAPD, the co-insurance rate slightly increased from 21% (95% CI, 21%–21%) in 2024 to 24% (95% CI, 24%–24%) in 2025, with tight clustering around these rates indicating consistent pricing strategies among insurers (Figure 1D). Conversely, Tier 4 MAPD co-insurance rates decreased on average from 45% (95% CI, 44%–45%) to 37% (95% CI, 37%–38%) but exhibited wider variability (range 20%–50%), suggesting heterogeneity in insurer strategies for higher-cost medications. PDPs maintained stable co-insurance structures, indicating limited strategic shifts due to the previously established use of co-insurance.

### Formulary Coverage and Utilization Management Changes

Besides benefit design, Part D plans can utlize formulary restrictions and utilization management strategies—including Quantity Limits (QL), Prior Authorization (PA), and Step Therapy (ST)— to regulate medication access, particularly for higher-cost medications.^14,18^

In Figure 2A, the average number of covered medications per plan modestly decreased from 2024 to 2025 for both PDP and MAPD plans, suggesting more restrictive formulary coverage. MAPD plans consistently covered more drugs overall compared to PDPs in both years. The median number of drugs per plan subject to QL slightly increased from 2024 to 2025 across both PDP and MAPD plans, indicating increased use of QL restrictions. The total number of medications requiring PA or ST remained relatively stable across both plan types and years (Figure 2B).

**Figure 2.**
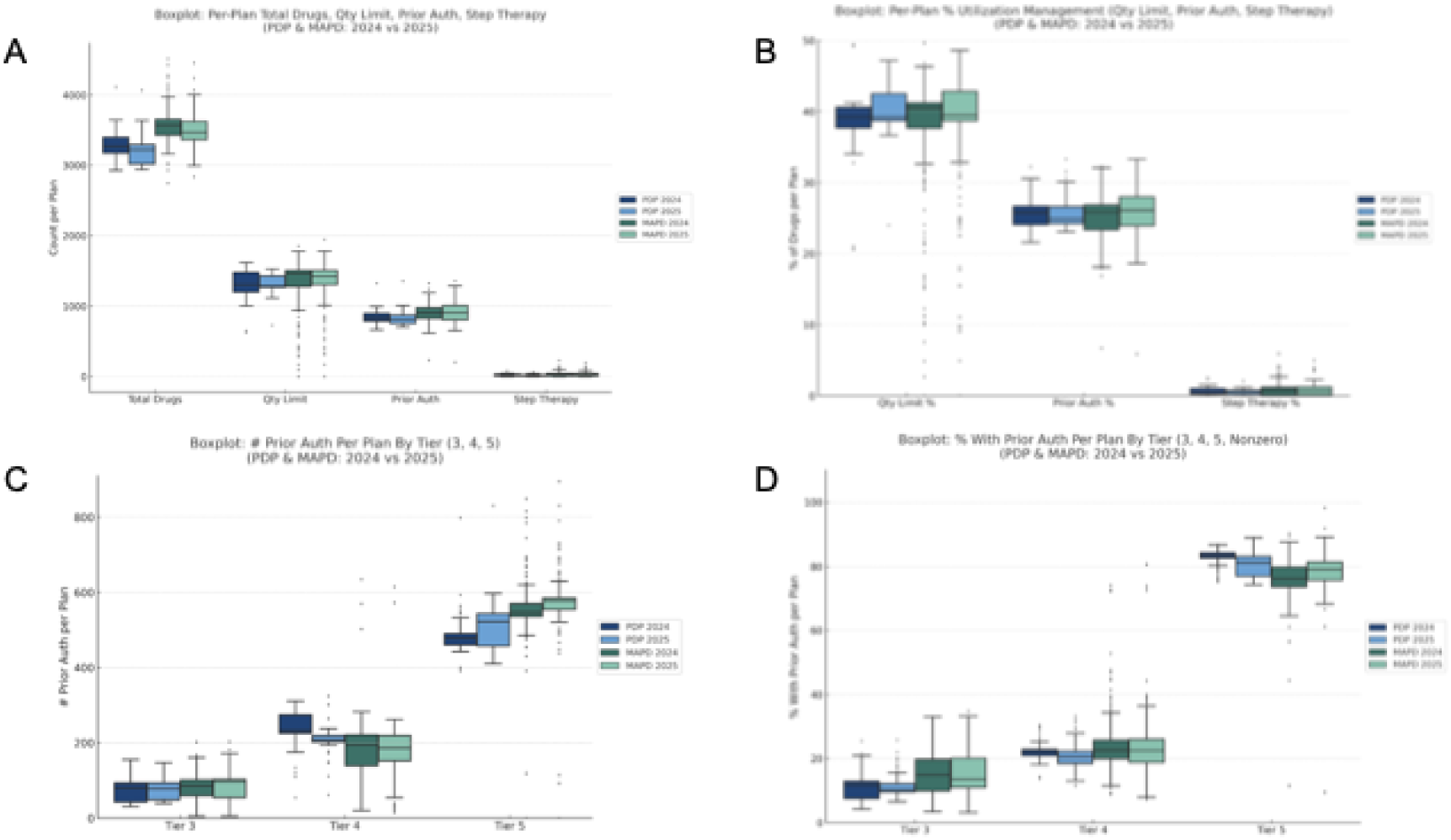
Changes in Medicare Part D Formulary Coverage and Utilization Management, 2024 vs 2025. **(A)** Total medications covered and number with Quantity Limits (QL), Prior Authorization (PA), and Step Therapy (ST) per plan type and year. **(B)** Percentage of covered drugs per plan subject to utilization management (QL, PA, ST). **(C)** Number of drugs per plan requiring PA across higher cost tiers (3–5). **(D)** Percentage of medications per plan requiring PA across tiers (3–5). Boxplots display median, interquartile ranges (IQR), whiskers (1.5× IQR), and outliers.

The utilization of PA across different tiers was further investigated (Figure 2C, 2D). From 2024 to 2025, the mean and median number of Tier 3 medications requiring PA remained stable in PDPs, with only a slight increase in MAPDs (mean: 85.1 to 88.7; median: 86 to 97). Interestingly, both PDPs and MAPD plans showed a slight reduction in the total number of Tier 4 medications requiring PA from 2024 to 2025, while the percentage of PA utilization within Tier 4 decreased modestly or remained consistent.

Tier 5 (Specialty) medications consistently have the highest PA rates, with the mean percentage in PDPs exceeding 80% in both years (83.2% in 2024, 80.2% in 2025) and similarly high rates in MAPD plans (75.7% in 2024, 77.6% in 2025). While the total number of Tier 5 medications requiring PA increased in both PDPs (mean: 480 to 510) and MAPD plans (mean: 552 to 569), the percentage of Tier 5 medications requiring PA remained consistently high and stable (narrow IQR). This highlights the continued stringent control placed on specialty medications. Additionally, the variability for both the number and percentage of medications requiring PA is greater in MAPD plans, indicating more plan-to-plan differences than PDPs.

### Coverage, Access, and Affordability of GLP-1 Receptor Agonists

GLP-1 receptor agonists (GLP-1s) are widely recommended for managing Type 2 diabetes,^19^ and their costs and financial impact on patients and healthcare systems have sparked extensive research and discussions.^20–23^ Thus, we evaluated the impact of Medicare prescription coverage changes under the IRA on GLP-1 coverage, patient accessibility, and affordability.

First, both PDPs and MAPD plans covered fewer GLP1-s in 2025 compared to 2024, with PDPs decreasing from an average of 4.84 GLP-1s covered (95% CI, 4.73 – 4.95) to 3.97 (95% CI, 3.85 – 4.10) and MAPD plans decreasing from 5.31 (95% CI, 5.28 – 5.34) to 4.68 (95% CI, 4.65 – 4.72). MAPD plans consistently offered broader coverage than PDPs. Importantly, all plans in 2025 applied PA requirements for certain covered GLP-1s, compared to approximately 84% of plans in 2024, potentially increasing administrative burdens and barriers to patient access (Table 2).

**Table 2.**
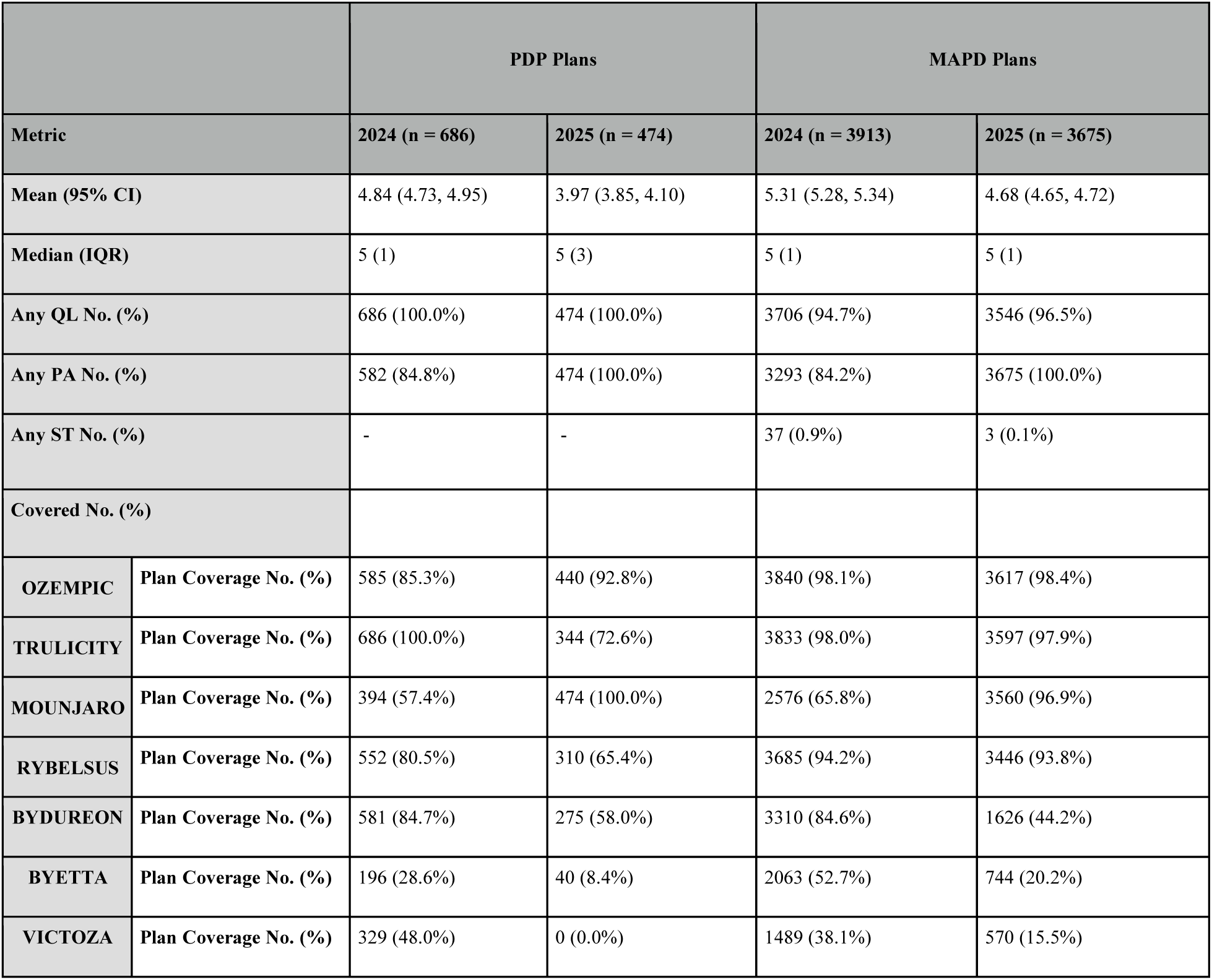
Medicare Part D Coverage and Utilization Management for GLP-1 Receptor Agonists (2024 vs 2025)

|  |  | PDP Plans |  | MAPD Plans |  |
| --- | --- | --- | --- | --- | --- |
| Metric |  | 2024 (n = 686) | 2025 (n = 474) | 2024 (n = 3913) | 2025 (n = 3675) |
| Mean (95% CI) |  | 4.84 (4.73, 4.95) | 3.97 (3.85, 4.10) | 5.31 (5.28, 5.34) | 4.68 (4.65, 4.72) |
| Median (IQR) |  | 5 (1) | 5 (3) | 5 (1) | 5 (1) |
| Any QL No. (%) |  | 686 (100.0%) | 474 (100.0%) | 3706 (94.7%) | 3546 (96.5%) |
| Any PA No. (%) |  | 582 (84.8%) | 474 (100.0%) | 3293 (84.2%) | 3675 (100.0%) |
| Any ST No. (%) |  | - | - | 37 (0.9%) | 3 (0.1%) |
| Covered No. (%) |  |  |  |  |  |
| OZEMPIC | Plan Coverage No. (%) | 585 (85.3%) | 440 (92.8%) | 3840 (98.1%) | 3617 (98.4%) |
| TRULICITY | Plan Coverage No. (%) | 686 (100.0%) | 344 (72.6%) | 3833 (98.0%) | 3597 (97.9%) |
| MOUNJARO | Plan Coverage No. (%) | 394 (57.4%) | 474 (100.0%) | 2576 (65.8%) | 3560 (96.9%) |
| RYBELSUS | Plan Coverage No. (%) | 552 (80.5%) | 310 (65.4%) | 3685 (94.2%) | 3446 (93.8%) |
| BYDUREON | Plan Coverage No. (%) | 581 (84.7%) | 275 (58.0%) | 3310 (84.6%) | 1626 (44.2%) |
| BYETTA | Plan Coverage No. (%) | 196 (28.6%) | 40 (8.4%) | 2063 (52.7%) | 744 (20.2%) |
| VICTOZA | Plan Coverage No. (%) | 329 (48.0%) | 0 (0.0%) | 1489 (38.1%) | 570 (15.5%) |

Four commonly prescribed GLP-1s (Ozempic, Mounjaro, Trulicity, Rybelsus) were further investigated regarding coverage and cost-sharing on plan-based and enrollment-based analyses (eTable 2). While plans tend to cover fewer GLP-1s in 2025, the coverage of Ozempic and Mounjaro has increased, with 17,714,559 (97.0%) PDP beneficiaries and 20,263,529 (98.6%) MAPD beneficiaries having access to Ozempic in 2025. The number of plans covering Mounjaro has increased dramatically from 394 (57.4%) to 474 (100.0%) within PDPs and 2,576 (65.8%) to 3,560 (96.9%) within MAPD plans, with all PDP beneficiaries and 19,025,316 (92.5%) MAPD beneficiaries having Mounjaro coverage. Previously, common options like Trulicity, Bydureon, Byetta, and Victoza showed substantially reduced coverage, highlighting a clear shift toward Ozempic and Mounjaro as preferred GLP-1 therapies.

Cost-sharing structures (co-payment and co-insurance) of four GLP-1s were further analyzed at the initial coverage stage through both plan-based and enrollment-weighted approaches. Under the co-payment (Figure 3A-B), both the median and mean patient OOP costs shifted upward in PDPs in 2025. Violin graphs show that 2025 PDPs exhibit a higher concentration of high co-payments for these GLP-1s, whereas PDPs in 2024 had a more even spread of OOP co-payments. Enrollment-weighted violin graphs show that nearly all patients face higher co-payments in 2025, with lower co-payments becoming rare. MAPD plans have comparatively modest changes in co-payment from 2024 to 2025 for these medications. Under co-insurance structures (Figure 3C-D), the enrollment-weighted analysis reveals a narrower and higher distribution of co-insurance rates for these GLP-1s in 2025 PDPs, indicating a consolidation of high co-insurance rates. This is even more apparent for MAPD plans in 2025, where average co-insurance payments are more concentrated at higher values compared to 2024, when a more even distribution is observed, with some concentration at lower costs. An exception to this trend was Mounjaro’s co-insurance payments, which decreased to the same level as other compared GLP-1s in 2025. This is likely a result of a large number of plans placing Mounjaro into Tier 3 coverage in 2025 as compared to higher tiers in 2024.

**Figure 3.**
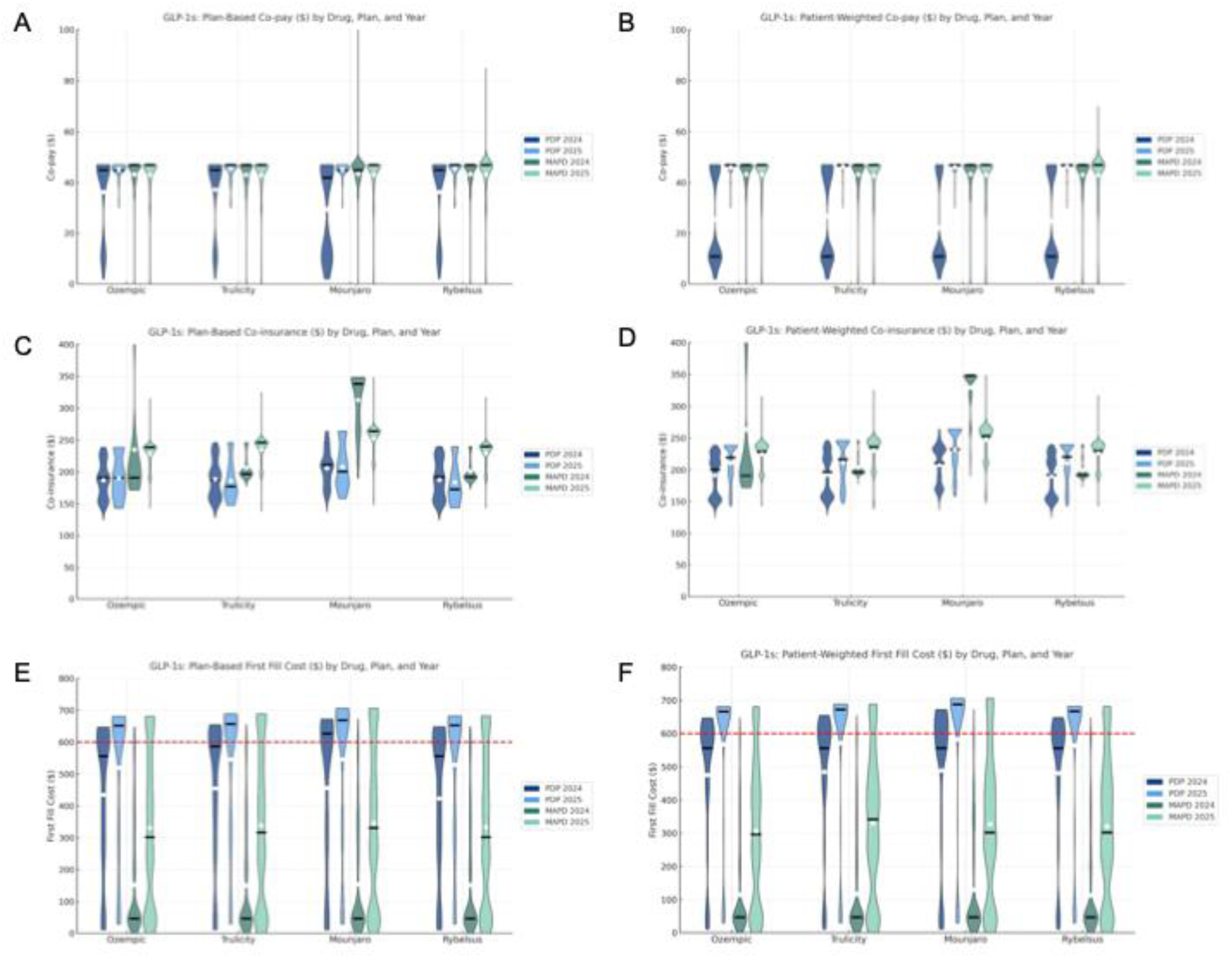
Cost-sharing and First-fill Cost for GLP-1s (2024 vs 2025) **(A)** Plan-level distribution of co-payment amounts for four commonly prescribed GLP-1s (Ozempic, Trulicity, Mounjaro, and Rybelsus) in PDP and MAPD plans, 2024 vs. 2025. **(B)** Enrollment-weighted distribution of co-payment amounts for four commonly prescribed GLP-1s for MAPD and PDP plans in 2024 and 2025. **(C)** Plan-level distribution of co-insurance amounts for GLP-1s across PDP and MAPD plans, 2024 vs. 2025. **(D)** Enrollment-weighted distribution of co-insurance amounts for GLP-1s across PDP and MAPD plans, 2024 vs. 2025. **(E)** Plan-based first-fill cost amount distribution for GLP-1s across PDP and MAPD plans, 2024 vs. 2025. **(F)** Enrollment-based first-fill cost amount distribution for GLP-1s across PDP and MAPD plans, 2024 vs. 2025. Black bars represent medians and white dots indicate means. The red dashed line in panels (E) and (F) marks the $600 threshold for first-fill costs.

Considering substantial deductible increases and the nature of Medicare prescription coverage phases, we further analyzed the financial burden of the initial (first-fill) GLP-1 prescription. We assumed patients filled the GLP-1 as their first medication in the year. The typical retail price of GLP-1s is approximately $900–$1,100 per 30-day fill. Most beneficiaries have a deductible, which is required to be fully paid, along with the initial stage of cost-sharing for the first-fill.

The mean and median first-fill costs dramatically increased in the plan and enrollment-based analysis of Ozempic, Mounjaro, Trulicity, and Rybelsus (Figures 3E and 3F, eTable 2). Furthermore, the first-fill costs for these GLP-1s under PDPs are consistently higher than with MAPD plans. For example, the first-fill cost of Ozempic was an average of $480.2 (95% CI, $523.3-$546.5) in 2024 and $565.2 (95% CI, $598.0-$631.9) in 2025 for PDPs, while for MAPD plans, the cost increased from $114.4 (95% CI, $81.4-$91.2) in 2024, to $305.0 (95% CI, $227.2-$245.1) in 2025 (enrollment-weighted analysis). This suggests that PDP beneficiaries encounter a higher financial barrier when attempting to obtain their first Ozempic prescription. Over 80% of PDP beneficiaries faced first-fill costs on these four GLP-1s exceeding $600 in 2025, compared to approximately 40-45% in 2024. Additionally, MAPD beneficiaries have experienced a nearly threefold rise in first-fill costs for these medications from 2024 to 2025. Overall, substantially increased first-fill medication costs will likely contribute to significant affordability challenges for beneficiaries initiating GLP-1 treatment this year.

## Discussion

The Medicare Part D policy reforms implemented in 2025 were intended to alleviate beneficiaries’ financial responsibility. However, our analysis suggests that Part D plans have strategically responded by adjusting their benefit designs—primarily through higher deductibles, increased use of co-insurance over co-payments in initial coverage, and narrowing formulary coverage.^24^ These strategies effectively shift more immediate prescription costs back onto beneficiaries, potentially offsetting some intended benefits of IRA provisions. A recent report shows PDPs increased co-insurance coverage utilization since 2020, while MAPD plans co-payment utilization remained stable from 2020 to 2024.^15^ Our analysis found a break in this stability when the use of co-insurance coverage in MAPD plans increased from 6.3% in 2024 to 38.1% in 2025 for Tier 3 medications, and from 19.7% in 2024 to 63.3% in 2025 for Tier 4 medications. Overall, MAPD plans are undergoing more drastic change compared to PDPs. More MAPD beneficiaries will now face increased financial uncertainty and higher deductibles for the first time, while PDP beneficiaries have consistently faced this challenge over the years.

For GLP-1s, our analysis revealed more concentrated coverage around specific medications, such as Ozempic and Mounjaro. Prior Authorization has become a commonplace requirement for most GLP-1s, which creates additional administrative burdens and potential delays in patient access.^4^ A potential benefit of this requirement is ensuring that GLP-1s indicated for diabetes are used only for patients with diabetes, eliminating GLP-1 shortages seen in 2023-2024 due to off-label use for weight loss (given poor coverage of weight-loss indicated GLP-1s within Medicare). Furthermore, financial analyses demonstrated dramatic increases in patient cost-sharing for both first-fills and subsequent fills of common GLP-1s due to significantly elevated deductibles and a substantial increase using coinsurance in initial coverage.^25,26^

In 2025, Medicare also launched Medicare Prescription Payment Plan (MPPP), allowing beneficiaries to enroll in the program and pay OOP cost in a capped monthly payment.^27,28^ This program aimed to distribute the cost of large lump sum payments at the pharmacy into monthly payments directly to the insurance. Formulas used to calculate these monthly payments are based on the $2,000 TrOOP cap, incoming OOP costs, and months remaining in the calendar year (eFigure 2). Costs more than $600 will trigger ‘likely to benefit’ notices in pharmacies to alert patients about this option. Around 13% of MAPD and 80% of PDP beneficiaries initiating GLP-1s would exceed the $600 threshold on their first-fill, making them prime candidates for this new payment option. A recent report stated that only 0.32% (n = 179,000) of beneficiaries enrolled in MPPP during the first two months of 2025, suggesting low overall program utilization.^29^ Furthermore, since the monthly payment amount is also determined by the timing of enrollment in MPPP, those who enroll later in the calendar year may not receive the full benefits of MPPP, given there are fewer months left for cost distribution. Expanding awareness and utilization of MPPP among healthcare providers, pharmacists, and beneficiaries could mitigate some immediate financial barriers and support sustained medication adherence.^30,31^

On an annual basis, the IRA capped OOP expenses at $2,000 for Medicare beneficiaries in 2025, down significantly from $8000 in 2024.^8^ Our analysis shows that Part D insurers countered these changes through several different mechanisms which shifted cost-sharing back onto Medicare beneficiaries. As a result, Medicare beneficiaries are not able to appreciate the annualized savings enacted by IRA as the first-fill cost of any Tier 3-5 drug can be prohibitively expensive, leaving patients unable to start many first-line guideline-directed therapies. MPPP attempted to address this issue; however, it has been underutilized thus far. In the 2026 calendar year, Medicare prescription coverage will continue to follow its current structure with small changes. It will be important to consider the impact of the Medicare Drug Price Negotiation Program as the first 10 medications in the program will have their prices reduced on January 1, 2026.^32,33^ Further analysis is needed to understand the full impact on the true affordability of these medications.

## Limitations

The study’s limitations include that no claims-level data was applied and analyzed to test the real-world influence of the new plan benefit design; we only focused on GLP-1s as an example, but similar affordability and access issues likely extend to other costly medication classes, including oncology treatments, specialty medications, and biologics, warranting further comprehensive evaluation. Additionally, the analysis was limited to commercial MAPD and PDP plans, excluding Special Needs Plans (SNP), Programs of All-Inclusive Care for the Elderly (PACE), Medicare-Medicaid plans, and employer-sponsored Medicare plans. Beneficiaries enrolled in these specialized plans may experience distinct benefit design responses to IRA policy changes, and thus warrant separate investigation.

## Conclusions

The study found that Medicare beneficiaries faced a higher deductible, more coinsurance coverage resulting in higher fill costs, and narrower formulary options in 2025. The IRA aims to alleviate patients’ cost burden, while Part D plans have implemented adaptive benefit strategies to maximize patient financial responsibility, which has created new barriers for medication affordability and access. Future research should incorporate claims data to assess how these benefit design shifts influence patients’ medication adherence and overall healthcare outcomes.

## Data Availability

All data produced in the present study are available upon reasonable request to the authors

## References

1. Medicare Monthly Enrollment. CMS Data. Accessed 18 June, 2025. https://data.cms.gov/summary-statistics-on-beneficiary-enrollment/medicare-and-medicaid-reports/medicare-monthly-enrollment

2. Medicare 101. KFF. Accessed 23 June, 2025. https://www.kff.org/health-policy-101-medicare/?entry=table-of-contents-who-is-covered-by-medicare

3. Trish E, Van Nuys K, Wu J, Desai NR. The Inflation Reduction Act and Patient Costs for Drugs to Treat Heart Failure. JAMA Netw Open. Oct 1 2024;7(10):e2441915. doi:10.1001/jamanetworkopen.2024.41915

4. Young GM, Bansal K, Riello RJ, 3rd, et al. Medicare Coverage and Patient Out-of-Pocket Costs for Cardiovascular-Kidney-Metabolic Medications. JAMA Netw Open. May 1 2024;7(5):e2412437. doi:10.1001/jamanetworkopen.2024.12437

5. Buttorff C, James HO, Sorbero ME, Reid RO. Medicare Part D insulin coverage: formulary strategies amid policy headwinds. Health Aff Sch. Apr 2025;3(4):qxaf042. doi:10.1093/haschl/qxaf042

6. Tarazi W, Finegold K, Sheingold S, De Lew N, Sommers B. Prescription Drug Affordability among Medicare Beneficiaries. *Office of the Assistant Secretary for Planning and Evaluation*, US Department of Health and Human Services. 2022;(Issue Brief No. HP-2022-03)

7. He Q, Silverman CL, Park C, Tiu GF, Ng BP. Prescription drug coverage satisfaction, cost-reducing behavior, and medication nonadherence among Medicare beneficiaries with type 2 diabetes. Journal of Managed Care & Specialty Pharmacy. 2021;27(6):696–705. doi:10.18553/jmcp.2021.27.6.696

8. Medicare Part D Improvements. CMS. Accessed 23 June, 2025. https://www.cms.gov/priorities/medicare-prescription-drug-affordability/overview/medicare-part-d-improvements

9. Sachs RE, Frank RG. Assessing the Effect of the Medicare Part D Redesign. JAMA. Feb 25 2025;333(8):663–664. doi:10.1001/jama.2024.25827

10. Mein SA, Tale A, Rice MB, Narasimmaraj PR, Wadhera RK. Out-of-Pocket Prescription Drug Savings for Medicare Beneficiaries with Asthma and COPD Under the Inflation Reduction Act. J Gen Intern Med. Apr 2025;40(5):1141–1149. doi:10.1007/s11606-024-09063-4

11. Dusetzina SB. Improving the Affordability of Prescription Drugs for Medicare Beneficiaries. JAMA Intern Med. Mar 1 2025;185(3):260–261. doi:10.1001/jamainternmed.2024.6915

12. Part D payment system. MedPAC. Accessed 23 June, 2025. https://www.medpac.gov/wp-content/uploads/2024/10/MedPAC_Payment_Basics_24_PartD_FINAL_SEC.pdf

13. Cai CL, Kesselheim AS, Bhaskar A, Rome BN. Insurer Exits After the Inflation Reduction Act Part D Redesign. JAMA. May 14 2025;doi:10.1001/jama.2025.7289

14. Jiang C, Nipp RD, Hong AS, et al. Prior Authorization, Quantity Limits, and Costs for Varenicline in Medicare. JAMA Netw Open. Mar 3 2025;8(3):e250008. doi:10.1001/jamanetworkopen.2025.0008

15. Trish E, Blaylock B, Van Nuys K. Cost Sharing for Preferred Branded Drugs in Medicare Part D. JAMA. Feb 14 2025;333(13):1170–2. doi:10.1001/jama.2024.28092

16. Lakdawalla D, Li M. Association of Drug Rebates and Competition With Out-of-Pocket Coinsurance in Medicare Part D, 2014 to 2018. JAMA Netw Open. May 3 2021;4(5):e219030. doi:10.1001/jamanetworkopen.2021.9030

17. Fendrick AM, Axelsen K. Medicare Reforms Necessitate More Formulary Oversight. Health Affairs Forefront. January 23 2025; doi:10.1377/forefront.20250121.933218

18. Klebanoff MJ, Li P, Lin JK, Doshi JA. Formulary Coverage of Brand-Name Adalimumab and Biosimilars Across Medicare Part D Plans. JAMA. Jul 2 2024;332(1):74–76. doi:10.1001/jama.2024.8917

19. Kalyani RR, Neumiller JJ, Maruthur NM, Wexler DJ. Diagnosis and Treatment of Type 2 Diabetes in Adults: A Review. JAMA. Jun 23 2025; doi:10.1001/jama.2025.5956

20. Tsipas S, Khan T, Loustalot F, Myftari K, Wozniak G. Spending on Glucagon-Like Peptide-1 Receptor Agonists Among US Adults. JAMA Network Open. 2025;8(4):e252964–e252964. doi:10.1001/jamanetworkopen.2025.2964

21. Hwang JH, Laiteerapong N, Huang ES, Mozaffarian D, Fendrick AM, Kim DD. Fiscal Impact of Expanded Medicare Coverage for GLP-1 Receptor Agonists to Treat Obesity. JAMA Health Forum. Apr 4 2025;6(4):e250905. doi:10.1001/jamahealthforum.2025.0905

22. Zhang YW, Lin NP, Guo X, Szabo-Fresnais N, Ortoleva PJ, Chou DH. Omniligase-1-Mediated Phage-Peptide Library Modification and Insulin Engineering. ACS Chem Biol. Feb 16 2024;19(2):506–515. doi:10.1021/acschembio.3c00685

23. DeJong C, Masuda C, Chen R, Kazi DS, Dudley RA, Tseng CW. Out-of-Pocket Costs for Novel Guideline-Directed Diabetes Therapies Under Medicare Part D. JAMA Intern Med. Dec 1 2020;180(12):1696–1699. doi:10.1001/jamainternmed.2020.2922

24. Medicare Part D in 2025: A First Look at Prescription Drug Plan Availability, Premiums, and Cost Sharing. KFF. Accessed 23 June, 2025. https://www.kff.org/medicare/issue-brief/medicare-part-d-in-2025-a-first-look-at-prescription-drug-plan-availability-premiums-and-cost-sharing/

25. Barthold D, Li J, Basu A. Patient Out-of-Pocket Costs for Type 2 Diabetes Medications When Aging Into Medicare. JAMA Network Open. 2024;7(7):e2420724–e2420724. doi:10.1001/jamanetworkopen.2024.20724

26. Trish E, Kaiser K, Joyce G. Association of Out-of-Pocket Spending With Insulin Adherence in Medicare Part D. JAMA Netw Open. Jan 4 2021;4(1):e2033988. doi:10.1001/jamanetworkopen.2020.33988

27. Medicare Prescription Payment Plan. CMS. Accessed 23 June, 2025. https://www.cms.gov/medicare/health-drug-plans/medicare-prescription-payment-plan

28. Doshi JA, Li P, Klebanoff MJ, Lin JK. Inflation Reduction Act Provisions and Medicare Part D Out-of-Pocket Costs for Specialty Drugs. JAMA Health Forum. May 2 2025;6(5):e251387. doi:10.1001/jamahealthforum.2025.1387

29. Duke D, Cline M, Liner D. MedIntel Insights: Early look at Medicare Prescription Payment Plan enrollment. Accessed 2 July, 2025. https://www.milliman.com/en/insight/medintel-insights-early-look-m3p-enrollment

30. Lalani HS, Hwang CS, Kesselheim AS, Rome BN. Strategies to Help Patients Navigate High Prescription Drug Costs. JAMA. Nov 26 2024;332(20):1741–1749. doi:10.1001/jama.2024.17275

31. Doshi JA, Li P, Asthana S, Lin JK. Reducing Medicare Part D Out-of-Pocket Costs for Specialty Oral Anticancer Drugs Under the Inflation Reduction Act: Highlighting the Benefits of Enrolling in the Medicare Prescription Payment Plan. JCO Oncol Pract. Apr 25 2025:OP2400937. doi:10.1200/OP-24-00937

32. Medicare Drug Price Negotiation Program. CMS. Accessed 23 June, 2025. https://www.cms.gov/priorities/medicare-prescription-drug-affordability/overview/medicare-drug-price-negotiation-programs

33. Patterson JA, Wagner TD, O’Brien JM, Campbell JD. Medicare Part D Coverage of Drugs Selected for the Drug Price Negotiation Program. JAMA Health Forum. Feb 2 2024;5(2):e235237. doi:10.1001/jamahealthforum.2023.5237

