## Supplements for "Assessing the Impact of the Inflation Reduction Act on Medicare Prescription Drug Coverage"

Supplement eTable 1. Changes in Medicare Part D Cost-Sharing Structures, 2024 vs 2025
Supplement eTable 2. Enrollment-weighted Coverage, Cost-sharing and First-fill Cost for GLP-1s (2024 vs 2025)
Supplement eFigure 1. Structure change of Medicare Part D in 2025 vs 2024
Supplement eFigure 2. Formulas for Medicare Prescription Payment Plan (MPPP)

### Supplement eTable 1. Changes in Medicare Part D Cost-Sharing Structures, 2024 vs 2025

| | PDP Plans | | MAPD Plans | |
|---|---|---|---|---|
| **Metric** | **2024 (n = 686)** | **2025 (n = 474)** | **2024 (n = 3913)** | **2025 (n = 3675)** |
| **Tier 3** | | | | |
| **Co-pay, No. (%)** | 276 (40.2 %) | 80 (16.9 %) | 3662 (93.6%) | 2269 (61.7%) |
| **Co-pay Mean (95% CI)** | 45.14 (44.76 – 45.52) | 45.29 (44.69 – 45.88) | 43.90 (43.66–44.14) | 43.82 (43.46–44.18) |
| **Co-pay Median (IQR)** | 47.00 (2.00) | 45.00 (2.00) | 47.00 (5.00) | 47.00 (2.00) |
| **Co-insurance, No. (%)** | 410 (59.8 %) | 394 (83.1 %) | 245 (6.3%) | 1401 (38.1%) |
| **Co- insurance Mean (95% CI)** | 0.20 (0.19 – 0.20 %) | 0.19 (0.19 – 0.20 %) | 0.21 (0.21–0.21) | 0.24 (0.24–0.24) |
| **Co- insurance Median (IQR)** | 0.20 (0.07) % | 0.19 (0.04) % | 0.20 (0.00) | 0.25 (0.02) |
| **Tier 4** | | | | |
| **Co-pay, No. (%)** | 2 (0.3 %) | 0 (0%) | 3135 (80.1%) | 1342 (36.5%) |
| **Co-pay Mean (95% CI)** | 96.50 (89.64–103.36) | — (N/A) | 96.22 (95.82–96.62) | 95.54 (94.74–96.35) |
| **Co-pay Median (IQR)** | 96.50 (3.50) | — (N/A) | 100.00 (5.00) | 100.00 (0.00) |
| **Co-insurance, No. (%)** | 684 (99.7%) | 474 (100%) | 772 (19.7%) | 2328 (63.3%) |
| **Co- insurance Mean (95% CI)** | 0.45 (0.45–0.46) | 0.43 (0.42–0.44) | 0.45 (0.44–0.45) | 0.37 (0.37–0.38) |
| **Co- insurance Median (IQR)** | 0.48 (0.09)% | 0.44 (0.12) | 0.50 (0.10) | 0.37 (0.25) |
| **Tier 5** | | | | |
| **Co-pay, No. (%)** | 0 (0%) | 0 (0%) | 0 (0%) | 3 (0.08%) |
| **Co-pay Mean (95% CI)** | — (N/A) | — (N/A) | — (N/A) | 0.00 (0.00–0.00) |
| **Co-pay Median (IQR)** | — (N/A) | — (N/A) | — (N/A) | 0.00 (0.00) |
| **Co-insurance, No. (%)** | 686 (100%) | 474 (100%) | 3873 (99.0%) | 3644 (99.2%) |
| **Co- insurance Mean (95% CI)** | 0.27 (0.27–0.28) | 0.27 (0.27–0.27) | 0.31 (0.31–0.31) | 0.30 (0.30–0.30) |
| **Co- insurance Median (IQR)** | 0.25 (0.05) | 0.25 (0.05) | 0.33 (0.03) | 0.30 (0.06) |

**Supplement eTable 2. Enrollment-weighted Coverage, Cost-sharing and First-fill Cost for GLP-1s (2024 vs 2025)**

| | | PDP Plans | | MAPD Plans | |
|---|---|---|---|---|---|
| **Metric** | | **2024 (n = 18,105,141)** | **2025 (n = 18,263,795)** | **2024 (n = 20,045,788)** | **2025 (n = 20,560,433)** |
| **OZEMPIC** | **Enrollment Coverage No. (%)** | 16,095,739 (88.9%) | 17,714,559 (97.0%) | 19,739,270 (98.5%) | 20,263,529 (98.6%) |
| | **Co-pay Mean [95%CI]** | 25.4 [14.8, 18.1] | 46.1 [46.5, 46.9] | 43.3 [42.8, 43.4] | 43.2 [42.8, 43.8] |
| | **Co-ins Mean [95%CI]** | 189.4 [189.1, 197.2] | 211.6 [214.1, 222.2] | 261.6 [356.6, 385.4] | 225.3 [212.0, 216.9] |
| | **First Fill Cost Mean [95%CI]** | 480.2 [523.3, 546.5] | 565.2 [598.0, 631.9] | 114.4 [81.4, 91.2] | 305.0 [227.2, 245.1] |
| | **First Fill Cost % > $600** | 41.6% | 82.2% | 0.6% | 13.1% |
| **TRULICITY** | **Enrollment Coverage No. (%)** | 17,720,489 (97.9%) | 14,015,753 (76.7%) | 18,729,026 (93.4%) | 19,287,723 (93.8%) |
| | **Co-pay Mean [95%CI]** | 27.3 [15.8, 19.2] | 46.4 [46.6, 47.0] | 43.1 [42.8, 43.4] | 42.8 [41.7, 42.9] |
| | **Co-ins Mean [95%CI]** | 192.4 [193.3, 201.2] | 213.2 [217.9, 227.7] | 204.3 [203.0, 207.9] | 231.0 [218.0, 223.3] |
| | **First Fill Cost Mean [95%CI]** | 488.1 [526.2, 549.3] | 568.1 [596.4, 636.0] | 114.8 [79.8, 87.7] | 321.9 [263.9, 280.2] |
| | **First Fill Cost % > $600** | 41.5% | 81.6% | 0.6% | 13.8% |
| **MOUNJARO** | **Enrollment Coverage No. (%)** | 11,464,579 (63.3%) | 18,263,795 (100.0%) | 13,991,578 (69.8%) | 19,025,316 (92.5%) |
| | **Co-pay Mean [95%CI]** | 21.7 [13.1, 16.8] | 46.1 [46.6, 46.9] | 43.5 [42.9, 43.8] | 43.0 [41.9, 43.0] |
| | **Co-ins Mean [95%CI]** | 208.9 [211.5, 220.6] | 232.1 [235.8, 245.2] | 330.0 [345.4, 347.2] | 247.8 [234.0, 239.3] |
| | **First Fill Cost Mean [95%CI]** | 485.6 [528.3, 561.3] | 586.6 [618.4, 651.3] | 127.7 [116.4, 137.7] | 328.5 [268.9, 287.8] |
| | **First Fill Cost % > $600** | 43.1% | 82.7% | 0.8% | 13.9% |
| **RYBELSUS** | **Enrollment Coverage No. (%)** | 16,004,030 (88.4%) | 13,466,517 (73.7%) | 18,308,766 (91.3%) | 18,754,566 (91.2%) |
| | **Co-pay Mean [95%CI]** | 25.4 [14.8, 18.4] | 46.4 [46.6, 47.0] | 43.2 [42.9, 43.5] | 42.9 [41.8, 42.9] |
| | **Co-ins Mean [95%CI]** | 190.3 [189.6, 198.4] | 210.0 [212.6, 222.1] | 199.1 [197.8, 202.5] | 226.0 [212.8, 217.9] |

| | | | | | |
|---|---|---|---|---|---|
| | **First Fill Cost Mean [95%CI]** | 479.9 [522.5, 546.5] | 560.0 [590.9, 631.4] | 115.0 [80.5, 89.2] | 320.6 [263.3, 280.2] |
| | **First Fill Cost % > $600** | 41.3% | 80.9% | 0.6% | 13.4% |

**Supplement eFigure 1. Structure change of Medicare Part D in 2025 vs 2024**

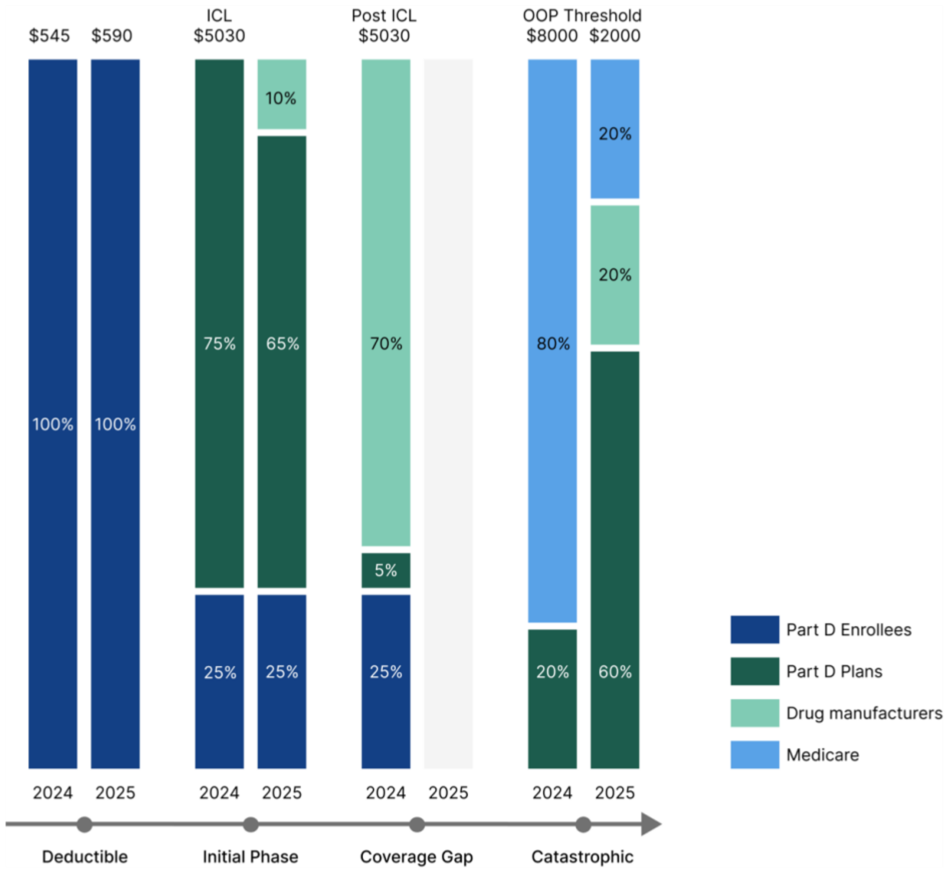

### Supplement eFigure 2. Formulas for Medicare Prescription Payment Plan (MPPP)

#### First Month Maximum Cap Bill Formula

$$\frac{\text{Annual OOP Threshold (\$2,000 in 2025)} - \text{any prescription costs the enrollee has already paid out of pocket that count toward TrOOP (True Out-of-Pocket)}}{\text{Number of months remaining in the year.}}$$

#### Subsequent Month Maximum Cap Bill Formula:

$$\frac{\text{Current M3P OOP Remaining Balance Due} + \text{New OOP Incurred Cost in the Month}}{\text{Number of months remaining in the year}}$$
